# COVID-19 Mitigation Practices and COVID-19 Rates in Schools: Report on Data from Florida, New York and Massachusetts

**DOI:** 10.1101/2021.05.19.21257467

**Authors:** Emily Oster, Rebecca Jack, Clare Halloran, John Schoof, Diana McLeod

## Abstract

This paper reports on the correlation of mitigation practices with staff and student COVID-19 case rates in Florida, New York, and Massachusetts during the 2020-2021 school year. We analyze data collected by the COVID-19 School Response Dashboard and focus on student density, ventilation upgrades, and masking. We find higher student COVID-19 rates in schools and districts with lower in-person density but no correlations in staff rates. Ventilation upgrades are correlated with lower rates in Florida but not in New York. We do not find any correlations with mask mandates. All rates are lower in the spring, after teacher vaccination is underway.

## I. Introduction

This paper uses data from the COVID-19 School Response Dashboard^1^ to report on mitigation practices in school and their correlation with staff and student COVID-19 rates over the 2020-2021 school year. The report focuses on three states with comprehensive data: Florida, New York and Massachusetts. The data are limited to public schools and districts.

We summarize several COVID-19 mitigation strategies adopted by school districts and correlate a subset of these mitigation strategies with COVID-19 case rates. The primary mitigation factors we focus on in the analysis are student in-person density levels, reported ventilation improvements and student and staff mask mandates. We differentiate between the pre- and post-vaccine availability period.

For students, COVID-19 rates are higher in schools and districts with lower in-person density. Staff rates are largely uncorrelated with student density, although in New York staff rates are higher in December in higher density districts. In the spring period, after vaccination is more widely available to teachers, we see little correlation between case rates and density in either direction in either population.

Ventilation improvements are correlated with lower COVID-19 rates in Florida in earlier periods but are not correlated in New York. These improvements vary, and we do not have detailed information on them. In the spring, there is little correlation between case rates and ventilation.

Mask mandates only vary across Florida. Some districts require masks for students and staff, some for staff only and some for neither. In terms of raw means, staff rates are higher in districts which do not have mask mandates for staff or students, although these differences are small. The differences are not significant in analyses which adjust for community rates. In all analyses, rates are similar for staff in districts with mask mandates for both students and staff versus those with staff-only mandates. Further, we do not see a correlation between mask mandates and COVID-19 rates among students in either adjusted or unadjusted analyses.

We caution that our analysis focuses only on correlations and it is challenging to make causal statements. In the case of masking in particular, we focus on *mandates* and not on actual behavior. Masking is likely correlated with mask mandates, but it is also likely that some individuals mask even in the absence of a mandate and that there is imperfect compliance even with a mandate. In addition, while we control for community rates, we do not control for community mitigation practices, which would also impact behavior and rates in schools.

This paper adds to our understanding of the relationship between COVID-19 mitigation and school safety in the US (Lessler et al., 2021; Varma et al., 2021; Zimmerman et al., 2021; van den Berg et al., 2021). We would emphasize that in general this literature suggests in-person school can be operated safely with appropriate mitigation, which typically includes universal masking. It would be premature to draw any alternative conclusions about this question based on this preliminary data.

This document is organized as follows: first, we describe the data collection approach for each state. Second, we summarize the analysis methods we use. Third, we summarize the mitigation approaches taken by each state. Finally, we discuss the relationship between mitigation and COVID-19 cases observed in the data.

## II. Data Collection Methods

For all states in our analysis (Florida, Massachusetts, and New York), we draw from data on public school- or district-level student enrollment (overall and in-person enrollment), COVID-19 case counts separated by students and teachers/staff, and mitigation practices. We collect these data directly from states, reviews of state websites, reviews of online school district reopening plans, and phone calls to districts without information online. We provide state-specific information about these data sources below.

### Florida

Florida data used for our analyses are publicly available online. COVID-19 case data are reported separately for students, teachers, and staff at the school-level each week and are available from the Florida Department of Health (https://floridahealthcovid19.gov/). Enrollment data are available at the school level from the Florida Department of Education (https://edstats.fldoe.org). These data are collected twice per school year, to measure fall and spring enrollment, and include the number of students who are enrolled in in-person, hybrid, and remote instruction. In-person teacher counts are from the school-level 2018-2019 NCES CCD, and the non-teacher staff counts are estimated from district-level counts.

Mitigation data come from systematic review of school district reopening plans performed by the COVID-19 School Dashboard team. Reviewers searched district plans to determine if each district reported using any of 13 different mitigation strategies across several areas. This included screening and testing, social distancing requirements, masking and ventilation requirements.

Reviewers then coded each school district as either using the given mitigation strategy or not (yes/no). In cases where school district plans did not include mitigation plans, reviewers emailed and made phone calls to school districts to clarify what mitigation practices were required.

### New York

Both COVID-19 case data and enrollment data are publicly posted online at the school-level from the New York COVID-19 Report Card (https://schoolcovidreportcard.health.ny.gov/#/home) on a weekly basis. The data were collected by the COVID-19 School Dashboard team on a biweekly basis by scraping the website. The state requires all schools to report COVID-19 test results separately for students and teachers/staff daily to the NY State Department of Health. These are reported as totals over the last 7 or 14 days.

Mitigation data were obtained through a systematic review of district plans following the same procedure as Florida.

### Massachusetts

Massachusetts district-level data on COVID-19 cases are reported weekly and publicly available from the Massachusetts Department of Elementary and Secondary Education (DESE) (https://www.doe.mass.edu/covid19/positive-cases/). The DESE also provides enrollment data to the COVID-19 School Response Dashboard on a weekly basis. These data include the number of students who are enrolled in in-person, hybrid, and remote instruction. In-person teacher and staff counts are from the district-level 2018-2019 NCES CCD.

No additional district specific mitigation data are used in this analysis as enrollment density is calculated from enrollment data and masking in schools is a universal requirement across the state. Evaluation of physical distancing practices in Massachusetts has been done previously with these data (van den Berg et al., 2021).

## III. Methods

We perform several analyses. First, we summarize mitigation strategies used by schools in each state. We report summary statistics weighting the data by enrollment. These summary data exclude fully remote districts. For Massachusetts we report these figures based on van den Berg et al. (2021).

Second, we show case rates for students and staff across all three states over time. These case rates are reported as daily case rates per 100,000 people. The case rates for students are cases as a share of in-person students.

Third, we explore correlations between mitigation strategies and COVID-19 case rates in students and staff. These analyses are limited to schools and districts with at least some in-person attendance and all analyses are weighted by total student enrollment.

We first show graphs of case rates over time by mitigation groups.

In-person student density is categorized into three groups based on the share of in-person student enrollment compared to total enrollment: 10%-49%, 50-79%, and 80% or more. Schools with in-person student density of less than 10% are defined as fully remote and excluded from the analysis. Schools in New York that are reported as “fully remote” are also excluded from the analysis regardless of reported density. This divides each state into roughly thirds, although Florida has greater in-person density in general. We note that density may be capturing various underlying policies, including distancing and hybrid school structures. We argue this is a useful measure, as it speaks directly to the ability of schools to open with close to full or full enrollment.

Ventilation is divided into two groups based on whether the districts reported ventilation improvements or not. Unfortunately, we do not have details about the specific improvements they may have made, and reported improvements may have been as simple as opening more windows.

For masking, we use data from Florida only (masking is universal in New York and Massachusetts). We report case rates in three groups: (1) Districts with mask mandates for all (22 districts, an average of 844,341 in-person students); (2) Districts with mask mandates only for staff (5 districts, 127,772 students); (3) Districts with no mask mandates (37 districts, 644,792 students). There are no districts with mandates for only students.

In addition to showing case rates over for students and staff in each state and mitigation group, we show effects adjusting for differences in community case rate and demographics.

To do this, we run regressions of the form below:

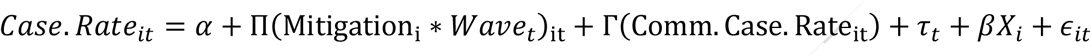

These regressions represent a standard “event-study” style analysis. We will graph coefficients over time for the groups, and report standard errors clustered by school districts.

In addition to graphical analyses, we will report effects aggregated over time in Appendix Tables, including a number of robustness checks and alternative functional forms.

## IV. Summary Statistics: Mitigation Strategies and Overall Case Rates

In Table 1, we report the population-weighted mitigation strategies used in each state. Student and staff masking requirements are universal in New York and Massachusetts, and occur in about half of Florida’s school districts. New York and Massachusetts have greater use of other mitigation practices as well, including a higher likelihood of student distancing requirements and more ventilation improvements in schools.

**Table 1.** Proportion of Students in Schools Utilizing Mitigation Practices in Florida, Massachusetts, and New York.

| **Mitigation Practice** | **Florida**<br>(n=1,453,464) | **Massachusetts**<br>(n=485,790) | **New York**<br>(n=1,223,327) |
| --- | --- | --- | --- |
| Student masks required | 0.54 | 1.00 | 1.00 |
| Staff masks required | 0.61 | 1.00 | 1.00 |
| Ventilation improvements | 0.64 | 0.80 | 0.92 |
| Students maintain 6 ft. | 0.79 | 0.72 | 0.98 |
| Students maintain 3 ft. | -- | 0.98 | -- |
| Student density (% of total enrollment) |  |  |  |
| Less than 10% | 0.02 | 0.17 | 0.04 |
| 10-49% | 0.25 | 0.12 | 0.39 |
| 50-79% | 0.47 | 0.41 | 0.33 |
| 80% or more | 0.26 | 0.30 | 0.24 |
| Staff testing prior to first day of school | 0.00 | -- | 0.02 |
| Daily at-home symptom screening | 0.90 | -- | 0.98 |
| Temperature check upon school entry | 0.75 | -- | 0.85 |
| Symptom check upon school entry | 0.64 | -- | 0.72 |
| Students stay in fixed cohorts during day | 0.53 | 0.88 | 0.87 |
| Groups restricted to under 25 people | 0.07 | 0.88 | 0.10 |
| Students stay in classroom and teachers rotate | 0.39 | -- | 0.69 |
| Move some class time outdoors | 0.29 | -- | 0.74 |

*Note:* This table reports student-weighted mitigation practices using coded mitigation practices from district reopening plans weighted by total student enrollment. Other than density, mitigation practices are summarized only for districts which are not virtual. Coded mitigation practices for FL and NY did not include 3 ft. distancing.

Student density ranges from less than 10% (which we define as remote) through greater than 90%. Florida has higher density schools than either Massachusetts or New York, on average.

Figure 1 reports overall case rates in each state among staff and students over the bi-weekly time periods in the data. The patterns are broadly consistent across all three states. Case rates increase over the fall, especially in December, and then fall rapidly in the spring. In New York and Massachusetts, student rates are lower than staff rates, although the converse is true in Florida. In the spring, staff rates fall more rapidly than student rates, likely reflecting vaccination among teachers.

**Figure 1.**
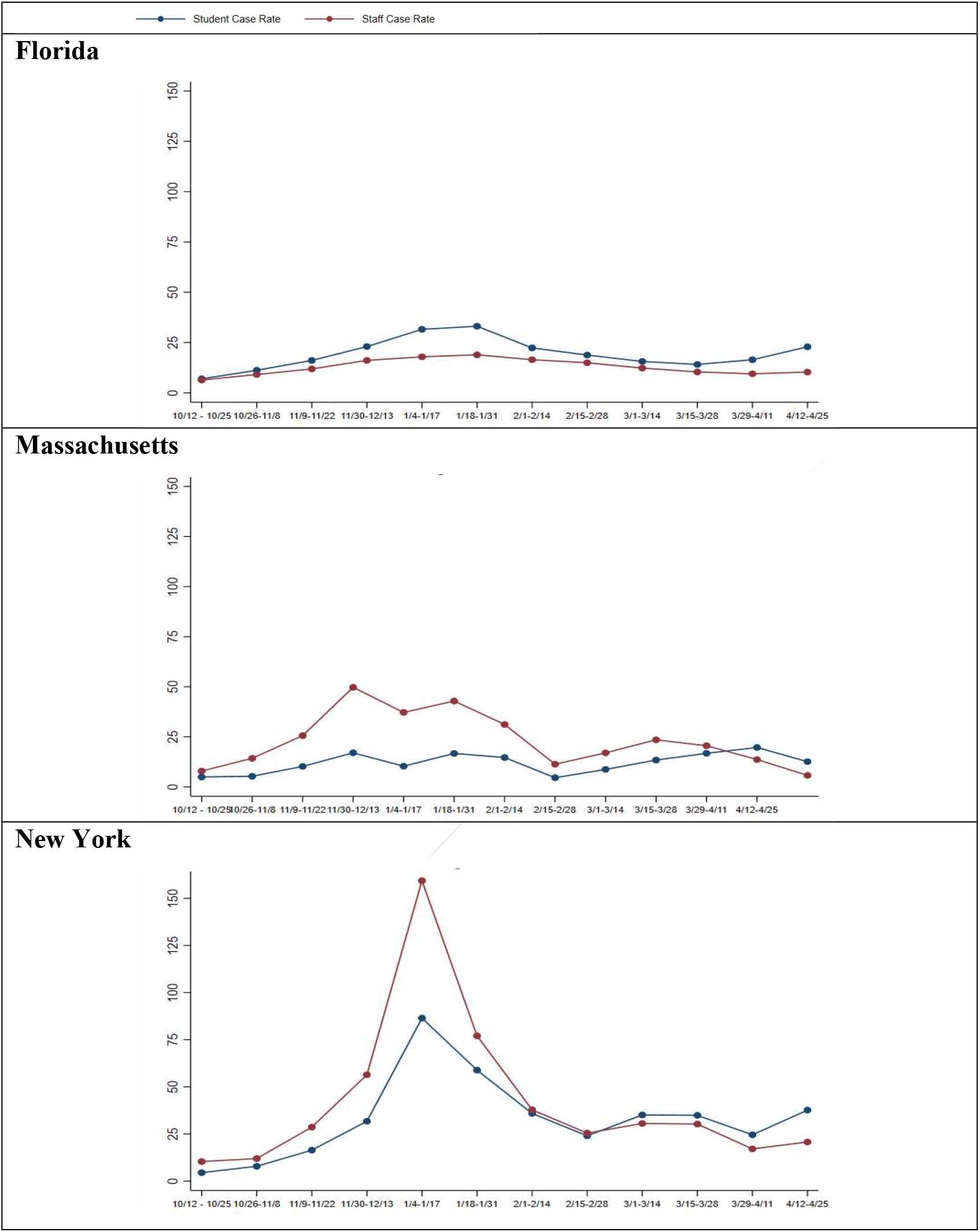
Cases Rates Among Staff and Students in Florida, Massachusetts, and New York. *Note*. These figures present the overall means of daily COVID-19 case rates per 100,000 in each state among staff and students over the bi-weekly time periods from October 2020 through April 2021. Mean daily case rate is calculated by group per biweekly wave in the data. Means do not control for community case rates or population demographics.

The case rates in New York are higher than in Massachusetts or Florida, which may reflect in part significantly higher testing among school participants in New York State.

## V. Correlation between Mitigation and Case Rates

We now turn to correlation between mitigation and case rates. The subsections below discuss in-person student density, ventilation improvements, and masking in turn.

In discussing these results, it is crucial to keep in mind that these data represent correlations and we cannot make strong causal claims. Notably, we expect school mitigation practices to reflect community mitigation practices: areas in which schools take more precautions will also take more precautions in other areas of society, making it difficult to attribute effects directly to the school behavior. In addition, these represent policies and may not necessarily reflect behavior. For example: masking may be widespread in schools even in the absence of a mandate. Conversely, even in the presence of a mask mandate, individuals may unmask in areas such as staff rooms, which may be higher risk.

### In-Person Student Density

Figure 2a shows student and staff means case rates by in-person student density. Figure 2b shows coefficients from regressions; the omitted category is the lowest density category (10% to 49%).

**Figure 2a.**
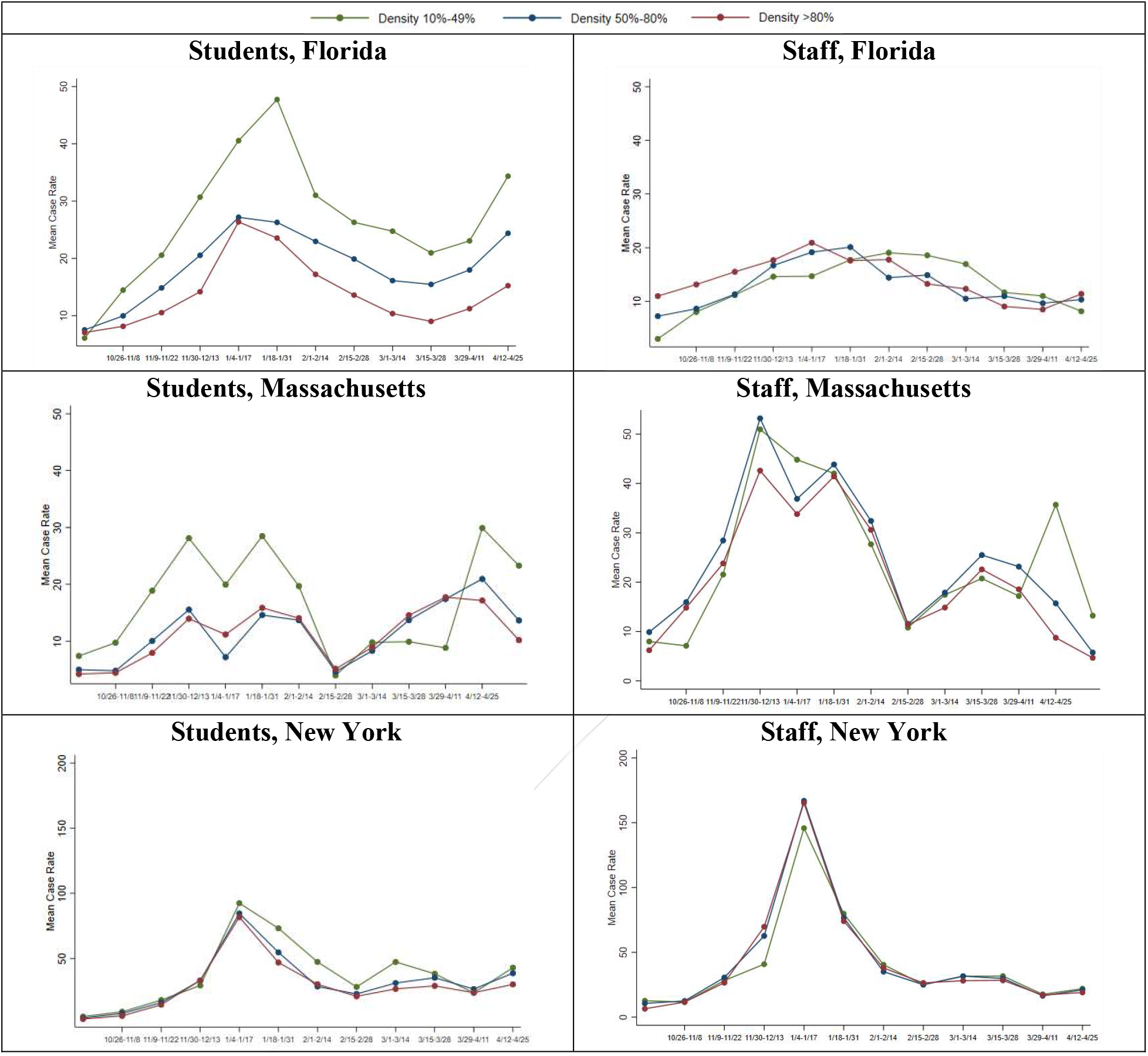
Mean Student and Staff Case Rates by Student Density in Florida, Massachusetts, and New York. *Note*. In-person student density is categorized into three groups based on the share of in-person student enrollment compared to total enrollment: 10%-49%, 50-79%, and 80% or more. Schools with in-person student density of less than 10% are defined as fully remote and excluded from the analysis. Case rates are reported as daily COVID-19 case rates per 100,000. Mean daily case rate is calculated by group per biweekly wave in the data. Means do not control for community case rates or population demographics.

**Figure 2b.**
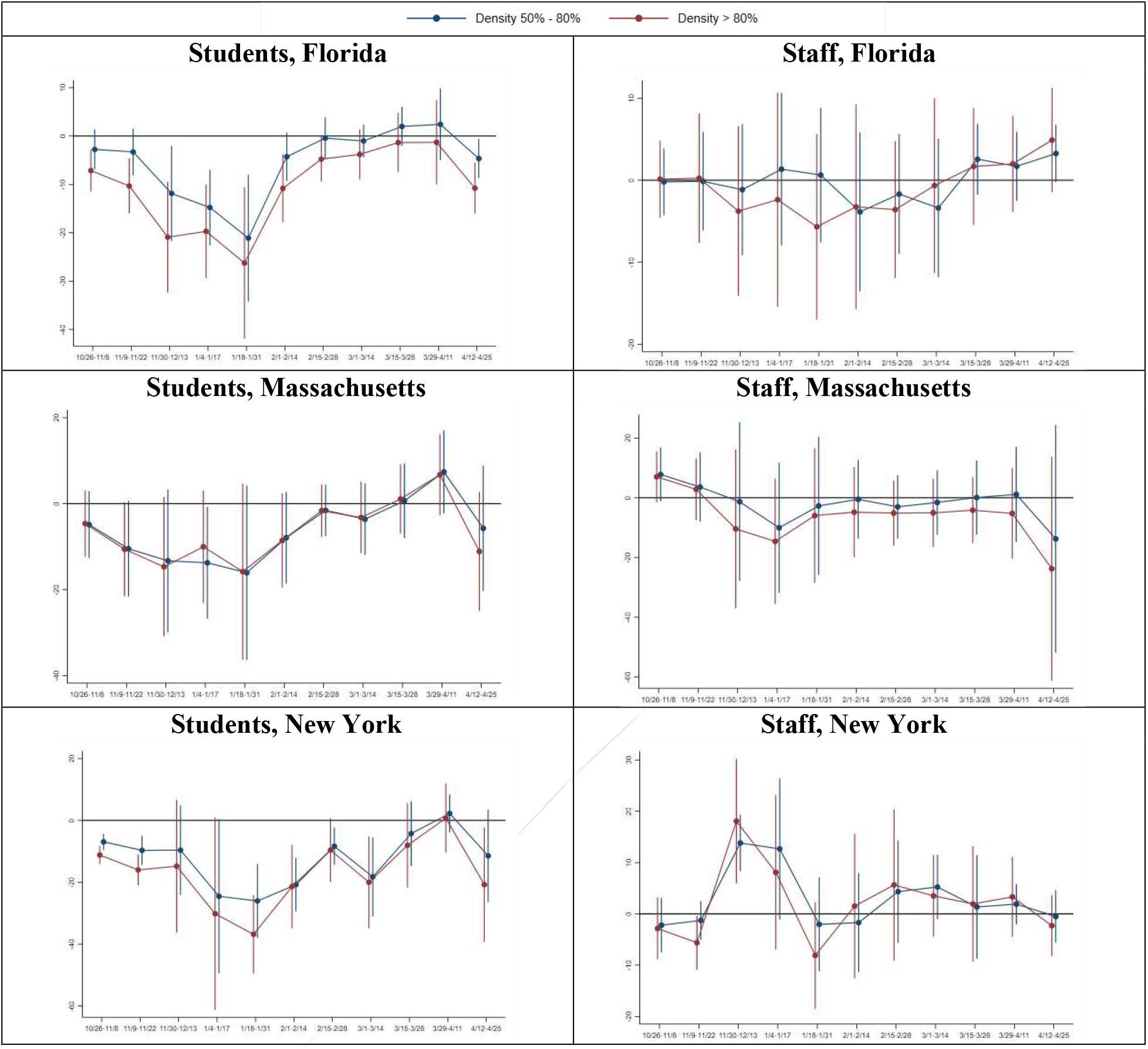
Regression Coefficients of Student and Staff Case Rates on Student Density in Florida, Massachusetts, and New York. *Note*. The regression coefficients are from regressions of student density groups interacted with each biweekly wave on student and staff case rates. Density of 10%-49% is used as the comparison group in all regressions. Regressions control for community case rates, time fixed effects, racial demographics and school level. The Florida regression also controls for masking groups (i.e. staff-only masks required and no masks required) and ventilation upgrades. The New York regression controls for ventilation upgrades. Regressions are weighted by total student enrollment and standard errors are clustered by school districts.

Across all three states, in both unadjusted means and coefficient estimates, a higher in-person student density is consistently associated with equal or *lower* school case rates among students. The lower rates among higher density districts are most pronounced in Florida, and most pronounced in the period of higher case rates in the winter.

In-person student density is uncorrelated with staff rates in either Florida or Massachusetts. In New York, we see higher case rates among staff in more dense districts in the pre-Christmas period.

The regression estimates in Table 2 confirm these results. Higher density is consistently associated with lower student case rates in all three states. The overall impacts on staff rates are insignificant in all three states.

**Table 2.** Regression Coefficient Estimates on Mitigation Practices for Student and Staff Case Rates, by State.

| Mitigation Practice,<br>by State | Main Model |  | No Community Case Rate<br>Controls |  | Poisson |  |
| --- | --- | --- | --- | --- | --- | --- |
|  | Student Rate | Staff Rate | Student Rate | Staff Rate | Student Rate | Staff Rate |
| **Florida** |  |  |  |  |  |  |
| Staff masks only required | 0.383<br>(2.131) | 2.395<br>(1.760) | -0.862<br>(2.186) | 1.165<br>(2.091) | 0.990<br>(0.111) | 1.171<br>(0.169) |
| No masks required | 2.318<br>(2.167) | 2.103<br>(2.094) | 2.396<br>(2.511) | 2.337<br>(2.550) | 1.116<br>(0.117) | 1.142<br>(0.164) |
| Ventilation improvements | -2.691<br>(2.297) | -2.661<br>(2.445) | -3.162<br>(2.360) | -4.088<br>(2.820) | 0.858<br>(0.0976) | 0.855<br>(0.136) |
| Student density |  |  |  |  |  |  |
| 50-79% | -6.136**<br>(2.436) | -0.103<br>(1.125) | -7.008**<br>(2.644) | -0.568<br>(1.486) | 0.773***<br>(0.0751) | 0.981<br>(0.0822) |
| 80% or more | -11.02***<br>(2.755) | -0.731<br>(1.818) | -12.84***<br>(2.838) | -1.944<br>(2.331) | 0.576***<br>(0.0705) | 0.916<br>(0.117) |
| Observations | 34,020 | 34,020 | 34,020 | 34,020 | 34,020 | 34,020 |
| **Massachusetts** |  |  |  |  |  |  |
| Student density |  |  |  |  |  |  |
| 50-79% | -5.729**<br>(2.508) | -1.416<br>(3.437) | -4.981*<br>(2.551) | 0.788<br>(3.854) | 0.655**<br>(0.112) | 0.941<br>(0.126) |
| 80% or more | -6.129**<br>(2.557) | -5.979*<br>(3.555) | -5.116**<br>(2.593) | -3.169<br>(3.693) | 0.627***<br>(0.108) | 0.777*<br>(0.110) |
| Observations | 3,080 | 3,080 | 3,080 | 3,080 | 3,080 | 3,080 |
| **New York** |  |  |  |  |  |  |
| Ventilation improvements | -1.915<br>(2.095) | -2.527<br>(2.466) | -2.686<br>(2.447) | -2.929<br>(2.905) | 0.938<br>(0.0542) | 0.943<br>(0.0542) |
| Student density |  |  |  |  |  |  |
| 50-79% | -10.79**<br>(5.437) | 1.970*<br>(1.010) | -9.316*<br>(5.036) | 3.182***<br>(1.094) | 0.708***<br>(0.0627) | 1.050*<br>(0.0295) |
| 80% or more | -15.56***<br>(5.364) | 0.972<br>(1.330) | -13.73***<br>(4.901) | 2.379<br>(1.604) | 0.628***<br>(0.0586) | 1.019<br>(0.0350) |
| Observations | 43,733 | 43,733 | 43,733 | 43,733 | 43,733 | 43,733 |

\*p-value < 0.05; \*\*p-value < 0.01; \*\*\*p-value < 0.001
*Note.* Regressions are weighted by total student enrollment and standard errors are clustered by school districts. Columns 1, 2, 5 and 6 include controls for community case rates, time fixed effects, racial demographics and school level. Columns 3 and 4 exclude community case rate controls but include all other controls. Columns 5 and 6 use a Poisson model and report coefficients as Incidence Rate Ratios. The omitted category for density is 10–49%; remote schools (defined as <10% density or reported as remote) are excluded from the analysis.

One interpretation of this finding for students is reflect differences in out of school activities. Places where density is lower are more likely to have students engaging with other groups (other childcare, family members, learning pods) when they are not in school. The extended groups may be a risk. A related interpretation is that families engaged in more extracurricular activities out of school when students had less in-school time.

### Ventilation

In New York and Florida, we are also able to look at ventilation improvements. Districts were coded as having ventilation improvements if their reopening plan mentioned any change in ventilation, including actions as simple as opening more windows.

Figure 3a shows staff and student rates by ventilation status in overall means. Figure 3b shows coefficients from regressions; the omitted category is the “no ventilation improvements” category.

**Figure 3a.**
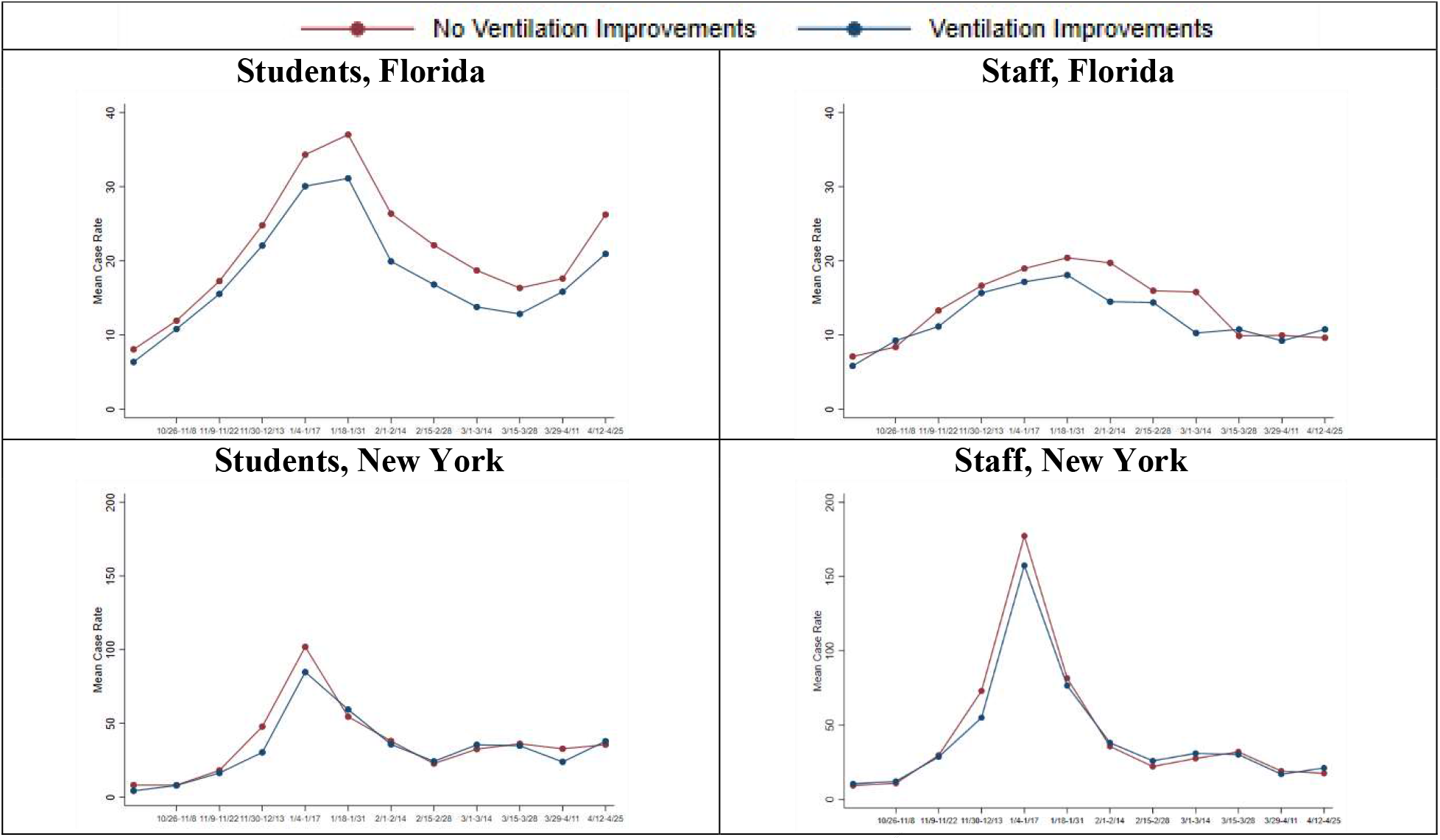
Mean Student and Staff Case Rates by Ventilation in Florida and New York. *Note*. Districts were coded as having ventilation improvements for any plan that mention changes in ventilation (e.g., opening more windows, installing new ventilation systems). Case rates are reported as daily COVID-19 case rates per 100,000. Mean daily case rate is calculated by group per biweekly wave in the data. Means do not control for community case rates or population demographics.

**Figure 3b.**
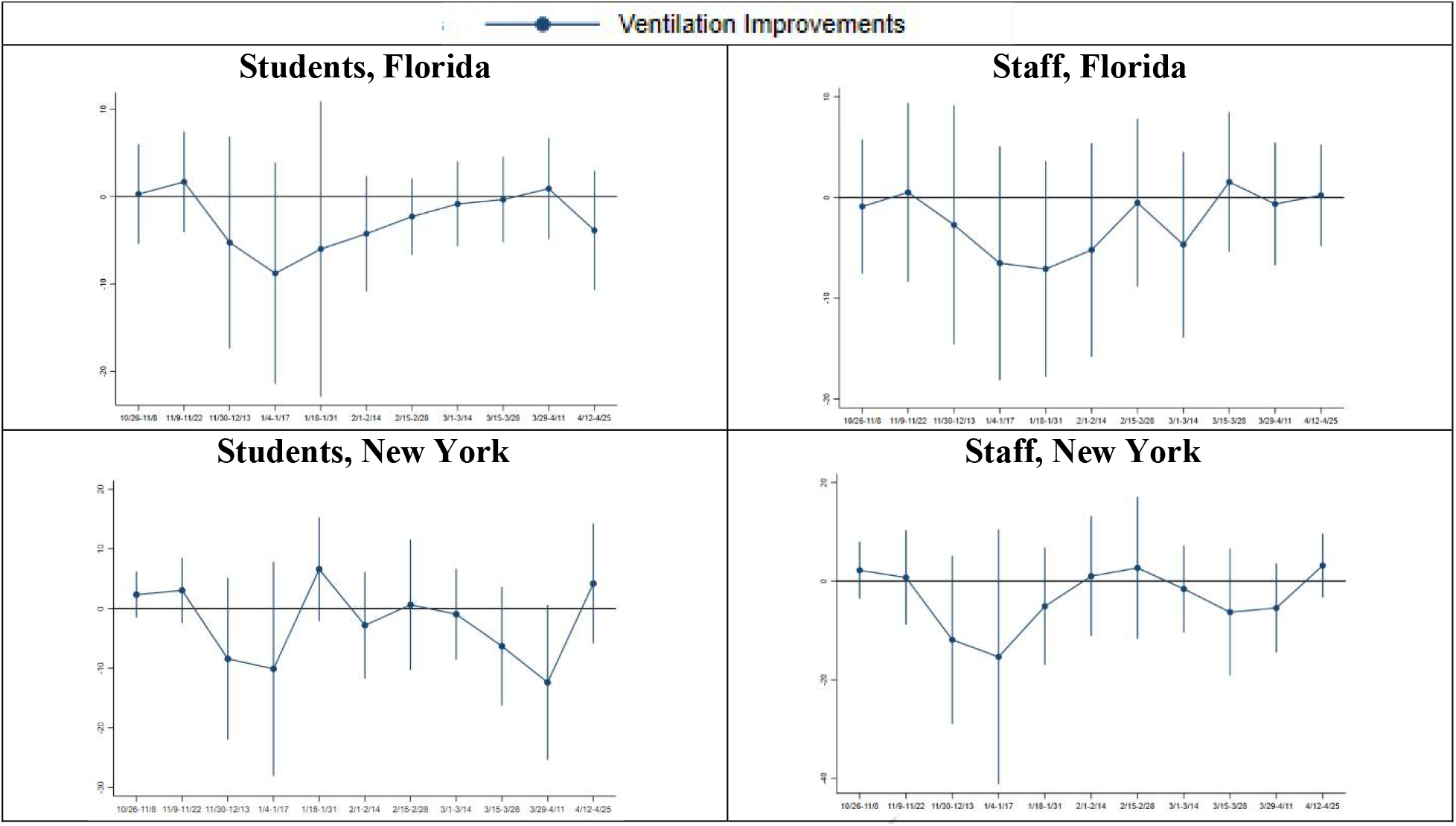
Regression Coefficients of Student and Staff Case Rates on Ventilation in Florida and New York. *Note*. The regression coefficients are from regressions of ventilation interacted with each wave on student and staff case rates. Regressions control for community case rates, time fixed effects, racial demographics, density groups, and school level. The Florida regression also controls for masking groups (i.e. staff-only masks required and no masks required). Regressions are weighted by total student enrollment and standard errors are clustered by school districts.

In unadjusted analyses, districts in Florida which do not report ventilation improvements have slightly higher case rates among staff. These differences do not persist in the adjusted analyses, or in the regressions in Table 2.

In New York we do not see evidence of variation in case rates based on ventilation.

It is important to note that given the wide range of possible ventilation improvements, it is difficult to be concrete here about which improvements might or might not matter.

### Masking

Figure 4a shows the overall case rates in the three masking groups in Florida (staff and student masks required, only staff masks required, no masks required); Figure 4b shows coefficients from regressions which adjust for case rates. Note that in Figure 4b the omitted category is “Mask Mandate for All” so the coefficients and significance are interpreted as relative to that group.

**Figure 4a.**
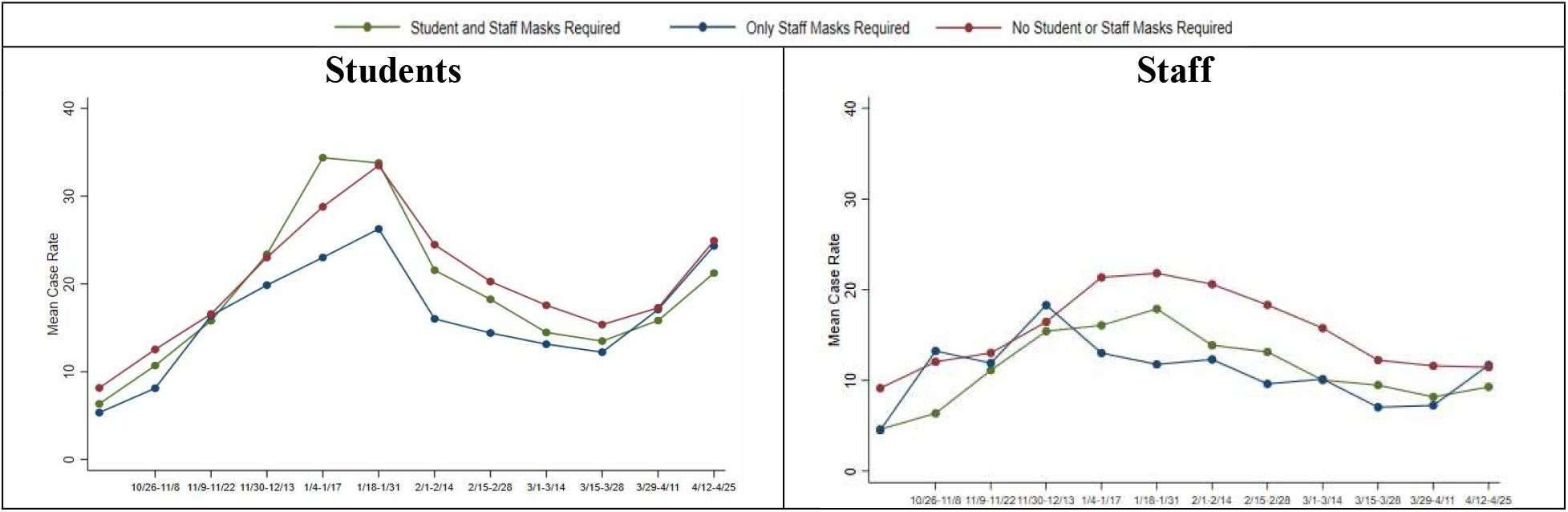
Mean Student and Staff Case Rates by Masking Requirements in Florida. *Note*. Florida masking practices are categorized into three groups: masks required for both students and staff, masks required for staff only, and no masks required for either students or staff. Case rates are reported as daily COVID-19 case rates per 100,000. Mean daily case rate is calculated by group per biweekly wave in the data. Means do not control for community case rates or population demographics.

**Figure 4b.**
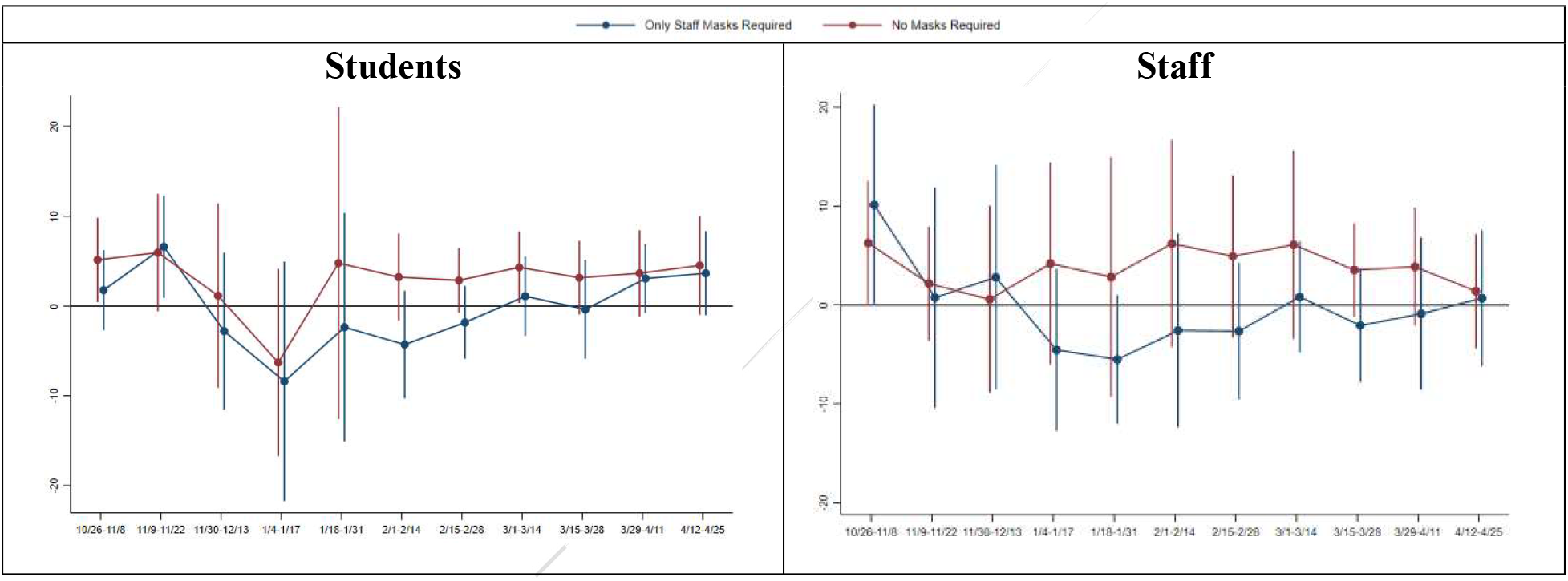
Regression Coefficients of Student and Staff Case Rates on Masking Requirements in Florida. *Note*. The regression coefficients are from regressions of masking groups (i.e. staff-only masks required and no masks required) interacted with each biweekly wave group on student and staff case rates. The comparison is masks required for both students and staff. Regressions control for community case rates, time fixed effects, racial demographics, density groups, ventilation upgrades, and school level. Regressions are weighted by total student enrollment and standard errors are clustered by school districts.

In Figure 4a we see higher staff COVID-19 rates in areas without mask mandates for either students or staff. Student COVID-19 rates do not appear to vary with mask mandates.

The results in Figure 4b are similar, although we find that the differences for staff are not significant once we adjust for community rates and other demographics. Community case rates appear to be higher in areas without mask mandates in schools, likely reflecting a lack of mask mandates in general.

The regressions in Table 2 are consistent with the figures; staff rates are slightly higher in areas without any mask mandates, but these results are not significant at conventional levels and are small.

It is important to note that this does not imply masks are ineffective, as these results focus only on masking in schools and do not take community behavior into consideration. Additionally, as noted above, we focus only on mask mandates and not actual masking behavior.

## VI. Discussion

We report here on correlations derived from data in the COVID-19 School Response Dashboard, which was combined with information on mitigation strategies recorded by school districts.

In all three states, we observe greater in-person density associated with lower case rates among students. One interpretation of this may reflect out-of-school activities. If student and teachers who are out of school engage in higher risk activities, more time in school could in principle be protective. This would be consistent with some existing literature (e.g. Mulligan, 2021; von Bismarck-Osten, Borusyak, & Schönberg, 2021). It is beyond the scope of these data to illustrate this mechanism directly.

Notably, in the spring period after vaccination is more widespread, we see very little variation across schools in staff or student case rates across any of these mitigation measures. Looking to the fall, this suggests that schools can operate safely in-person full-time.

This represents a preliminary analysis and carries limitations. First, we have comprehensive data for only three states, which are not representative of students across the U.S. as a whole. Second, there is variation in masking only in Florida, meaning that the data may be even less generalizable to all U.S. students.

Third, our data only represent cases among people associated with schools, *not* cases spread in schools. Careful contact tracing would be helpful in focusing on the latter, but is not widely available. Finally, we do not focus on possible community spread as a result of schools opening, which is a separate consideration and has been considered in other work (Courtemanche et al., 2021; Harris et al., 2021; Harell & Lieberman, 2021).

Future work with these data, and updated versions, may help shed more light on these issues. Given the challenges of virtual schooling (Diliberti & Kaufman, 2020), there is significant policy pressure to open fully in the fall and to establish the best approaches to doing so. The data here indicates higher density is at least not correlated with higher COVID-19 rates in schools. It may also suggest a more limited need for various mitigation measures in the fall, especially when staff and some older students are vaccinated.

## Supporting information

Raw Data for Paper

## Data Availability

Data is included with submission in .csv form.

## Footnotes

1 (https://covidschooldashboard.com/)

